# Genetic loss of *JAK1* and cutaneous HPV infection

**DOI:** 10.64898/2026.04.03.26350014

**Authors:** Shi-Qi Fan, Rong-Rong Wang, Roberto Colombo, Kai-Chen Tang, Jia-Wei Liu, Alessandro Pontoglio, Li-Li Zhang, Kai Li, Shi-Rui Han, Han Zhang, Xiao Bai, Xue Yu, Xiaerbati Habulieti, Ke-Qiang Liu, Yang Sun, Li-Wei Sun, Jia-Cheng Li, Hong Liu, Miao Sun, Zhi-Miao Lin, Fu-Ren Zhang, Dong-Lai Ma, Xue Zhang

## Abstract

**Background:** Human papillomaviruses (HPVs) pose a severe threat to global public health by driving nonmelanoma skin cancer (NMSC) and cervical cancer, with NMSC being one of the most common cancers worldwide. Epidermodysplasia verruciformis (EV) is an inborn error of immunity characterized by an increased susceptibility to persistent infection of cutaneous HPV and a high risk of NMSC. The genetic basis remains unknown in many patients with EV.

**Methods:** We collected four unrelated pedigrees with EV. Genetic analysis identified five variants in *JAK1* encoding the Janus kinase 1. *Ex vivo* models and patient-derived tissue were employed to evaluate the functional effects of *JAK1* variants and delineate the pathogenic mechanisms.

**Results:** We identified different variants in *JAK1* in four pedigrees with dominant EV. Genetic analysis revealed five novel variants in *JAK1*, three of which resulted in nonsense-mediated mRNA decay (NMD). Functional assays identified a decreased phosphorylation of the signal transducers and activators of transcription (STATs), impaired interferon responses, and defective T cell activation. Immune dysregulation in patients, characterized by a reduced CD4⁺/CD8⁺ T cell ratio, decreased CD8⁺ naïve T cell proportion, and accumulated memory T cells, implies impaired antiviral immunity against HPV.

**Conclusions:** Our findings confirm that *JAK1* loss-of-function (LOF) variants underlie susceptibility to cutaneous HPV infection. [Funded by the National Natural Science Foundation of China (81788101, 81230015, 82394420, and 82394423), the National Key Research and Development Program of China (2022YFC2703900), the CAMS Innovation Fund for Medical Sciences (2021-I2M-1-018), and the Regione Lombardia, Italy (Innovative Research Project 1137-2010)].

## INTRODUCTION

HPV is ubiquitous in the global population, which causes approximately 5% of all human cancers worldwide^1^. HPV-associated malignancies include nonmelanoma skin cancer (NMSC), cervical cancer, oropharyngeal cancer, anal cancer, and other tumor types. NMSC, including basal cell carcinoma (BCC) and cutaneous squamous cell carcinoma (cSCC), is the most common malignancy in the western world^2^. In addition, cervical cancer ranks the fourth most common cancer among women globally^3^. Annually, NMSC and cervical cancer account for nearly 15.3 million^4^ and 0.6 million^5^ new cases, respectively. Collectively, these diseases highlight the global health burden imposed by HPV infection. The cutaneous low-risk HPV is a potential facilitator of skin carcinogenesis and is detected in the majority of cSCC^6^. The link between carcinogenesis and viral infection has been known for a long period of time. HPV-associated cervical and cutaneous cancers exemplify this link. A vital role in determining the clinical consequence of an HPV infection is host-related factors, but unfortunately, the nature of this host-virus interplay is largely unknown.

Epidermodysplasia verruciformis (EV) (OMIM #226400), an inborn error of innate immunity, is the first described Mendelian disorder that links viral infection and cancer. Individuals with normal immunity resist the infection of low-risk HPVs and thus only manifest with asymptomatic infections or benign self-limiting papilloma^7^. In contrast, patients with EV are characterized by highly increased susceptibility to persistent cutaneous infection of a specific group of HPV, especially β-HPV. Such persistent HPV infection results in refractory, disseminated skin lesions manifesting as flat wart-like, common wart-like, seborrheic keratosis-like, and pityriasis versicolor-like lesions. Typically, these lesions appear in childhood or adolescence and persist, and confer a high risk of progression to NMSC^8^. About two-thirds of patients with EV will develop NMSC, especially on UV-exposed areas like the extremities, neck, and face^9^. Histopathological findings of EV include hyperkeratosis, parakeratosis, mild acanthosis, and koilocytes or vacuolated cells^10^.

EV can be roughly divided into typical EV, atypical EV, and acquired EV^11^. Both keratinocytes and T cells probably contribute to immunity to low-risk HPV. Phenotypes of typical EV are confined to the skin without extracutaneous manifestations. *TMC6*, *TMC8*, and *CIB1*, highly expressed in keratinocytes, are known causative genes for typical EV^12^. *ITGAL,* vital for T-cell skin homing, has been recently reported as a causative gene for typical EV^13^. Instead, patients with atypical EV usually develop EV in the context of other infectious diseases due to inborn errors of T cells or antigen-presenting cells (APC) immunity, and often display broader immunodeficiency manifestations^14^. Acquired EV is often found in organ transplantation and immunocompromised conditions such as Human Immunodeficiency Virus infection^15^.

JAK1 is critical for the JAK-STAT pathway, which could amplify cellular responses to interferon (IFN)^16^ and is essential for anti-viral immunity. Given the key role of JAK-STAT pathway hyperactivation in diverse disorders, JAK1 has emerged as a leading target for developing novel immunosuppressants. A single patient with biallelic loss-of-function (LOF) variants in *JAK1* has been described, presenting with immunodeficiency predominantly associated with mycobacterial infection^17^. In contrast, the heterozygous gain-of-function (GOF) variants in *JAK1* mainly account for immune dysregulation^18^ and autoimmune conditions^19^. In addition, owing to the wide range of signaling affected by JAK inhibitors, some adverse events, such as infection, are unpredictable^20^, especially the risk of NMSC^21^. No direct evidence of causality between *JAK1* variants and HPV susceptibility has been reported.

In this research, we collected four independent pedigrees of EV. All the patients harbor *JAK1* LOF variants (NM_002227.4, c.3258+1G>A in pedigree A; c.3169A>T in pedigree B; c.1730G>A and c.3103T>C in pedigree C; and c.425dupA in pedigree D). These *JAK1* variants caused impaired JAK-STAT pathway and defective anti-viral immunity in both T cells and keratinocytes. In summary, we identified *JAK1* as a novel causative gene for EV, which impairs the JAK-STAT pathway and anti-viral immunity, providing a mechanistic insight into HPV-specific immunity. These findings offer clinical insights to inform the prevention and treatment of NMSC and the application of JAK inhibitors, and also provide a unique model for investigating the interplay between HPV and host immunity.

## METHODS

### Study Oversight

Clinical information and samples were collected from the family trio, and informed consent was obtained. This study was approved by the Peking Union Medical College Institutional Review Board following the Declaration of Helsinki (Approval No: zs2024016).

### Genetic, Biochemical, and Functional analyses

Details of DNA extraction, whole exome/genome sequencing based on family trio (Trio-WES/WGS), variant screening, bioinformatic analyses and molecular modeling, cell culture, plasmids transfection, TOPO cloning, protein extraction and western blot, RNA extraction and reverse transcription quantitative PCR (RT-qPCR), flow cytometry, dual luciferase assay, viral DNA and antibody quantification, interleukins quantification, immunofluorescence staining, hematoxylin and eosin (H&E) staining, immunohistochemical staining, genotyping of HPV, cell proliferation assay, and single-cell RNA sequencing (scRNA-Seq), statistical analyses are described in **Supplementary Appendix 1**.

## RESULTS

### Patient characteristics

Four unrelated pedigrees with EV of different racial groups were collected from distinct geographical regions (**Fig. 1A**). These patients showed flat wart-like, common wart-like, seborrheic keratosis-like, or pityriasis versicolor-like skin lesions (**Fig. 1B, Fig. S1, and Fig. S2**), with typical vacuolated cells in the histopathological examination (**Fig. 1C**). Multiple EV-associated HPV types were subsequently determined by genotyping of DNA isolated from skin lesions. HPV types of pedigree A, C, and D **(Table S4)** and auxiliary examination results **(Table S5 to S8)** are summarized. The detailed clinical characteristics of patients from the four pedigrees are described in **Supplementary Appendix 2**. (To protect patient privacy, Supplementary Appendix 2 is available upon request from the corresponding author.)

(To protect patient privacy, access to Figure 1, Supplementary Figure 1, and Supplementary Figure 2 is available upon request from the corresponding author.)

**Figure 1.** Pedigrees, clinical characteristics, and histopathogenic staining. (Panel A) Four independent EV pedigrees were collected from distinct geographical regions. Probands were indicated by arrows. The solid symbols represent individuals who have phenotypes of EV. A red asterisk indicates that the individual has been infected with HBV. (Panel B) Patients from four pedigrees were all clinically diagnosed with EV. II-9 from the pedigree A manifests with an increasing number of flat and brown patches on his extremities, trunk, and face in sequence. II-1 and II-13 from the pedigree A display typical planar warts on their hands. III-1 from pedigree C has disseminated seborrheic-keratosis-like lesions on his face, neck, trunk, and upper limbs. IV-1 and IV-2 from the pedigree C developed multiple hyperplastic keratotic common-wart-like lesions and flat-wart-like lesions on the hands, upper limbs, and feet. IV-2 from the pedigree C has seborrheic-keratosis-like lesions on his face and neck. IV-2 from the pedigree D developed warts disseminated to his forehead, jaw, forearms, and perianal region. (Panel C) H&E staining of skin tissues from patients suggests the EV-specific vacuolated cells, indicating HPV infection. The black arrowhead indicates the vacuolated cells. The question mark indicates that phenotypes are not available.

(To protect patient privacy, access to Figure 1, Supplementary Figure 1, and Supplementary Figure 2 is available upon request from the corresponding author.)

**Supplementary Figure 1.** Clinical characteristics of pedigree. **A.** (Panel A) II-9, II-13, II-1, II-4, and III-12 from pedigree A manifested typical planar warts.

(To protect patient privacy, access to Figure 1, Supplementary Figure 1, and Supplementary Figure 2 is available upon request from the corresponding author.)

**Supplementary Figure 2.** Clinical characteristics of pedigree C and. **D.** (Panel A) III-1 from pedigree C manifested seborrheic keratosis-like lesions. These lesions developed and disseminated rapidly within one year. IV-2 from pedigree C manifested typical planar warts and common warts. Some lesions, exemplified by the one on the wrist, developed hyperpigmentation. His nails were also involved. (Panel B) IV-2 from pedigree D showed planar warts disseminated to the forehead, jaw, forearms, and perianal region. II-6 from the pedigree D had multiple seborrheic keratosis-like lesions and warts on the face, neck, and back. III-3 from pedigree D had multiple seborrheic keratosis-like lesions on his face, neck, and arms. The black arrow highlights a planar wart on his hand.

### Genetic testing revealed *JAK1* variants in different patients

Suspected variants in pedigrees A, C, and D were screened (**Fig. S3A**). No obvious region of loss of heterozygosity was observed in pedigrees A, C, and D, which again excludes consanguinity. All these independent pedigrees harbor *JAK1* variants (NM_002227.4, heterozygous variant c.3258+1G>A in pedigree A, heterozygous variant c.3169A>T in pedigree B, compound heterozygous variants c.1730G>A and c.3103T>C in pedigree C, and heterozygous variant c.425dupA in pedigree D) (**Table 1, Fig S3B**). *JAK1* has a high probability of being LOF intolerant (pLI) value of 1.000, and a low o/e value of 0.106 (90%CI 0.068-0.168), which indicates intolerance of LOF and possible pathogenic haploinsufficiency. All these variants were predicted to be pathogenic with an extremely low frequency in the population.

**Table 1.**
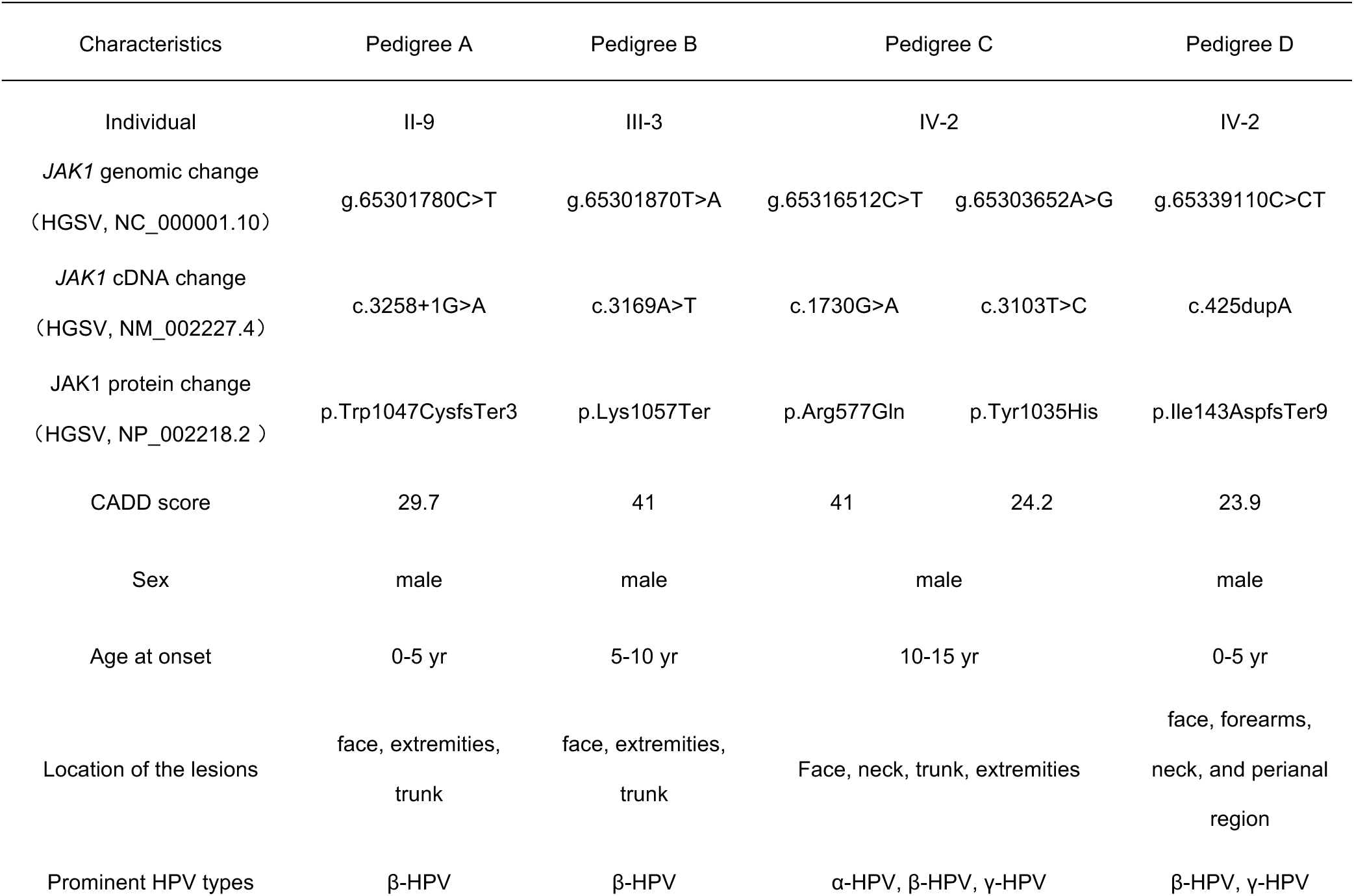
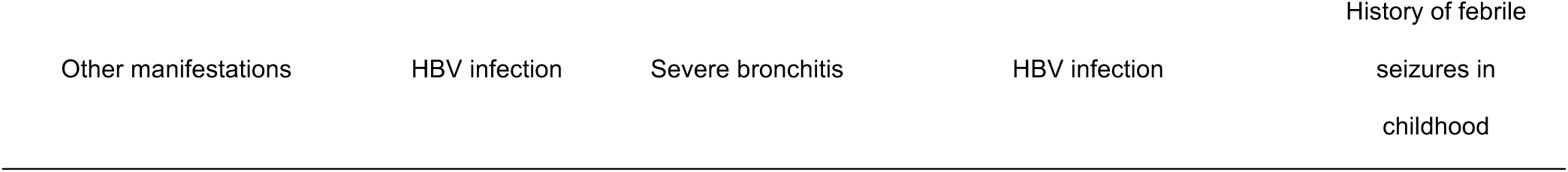
Basic information of the probands.

The c.3258+1G>A variant leads to an exon 23 skipping of *JAK1*, which was confirmed via the minigene assay and validated by RT-PCR followed by Sanger sequencing using peripheral blood samples (**Fig. 2C and 2D, Fig. S3D**). TOPO TA cloning suggested that the variant transcript was severely degraded (**Fig. S3E**). The amino acids in positions 577 and 1035 are highly conserved across species (**Fig. S3C**). All of these variants ranked a high CADD score (**Fig. 2A and Table 1**). Missense variants (c.1730G>A and c.3103T>C) were located in the region with high probability of mutation intolerance (**Fig. 2B and Fig. 2E**). The c.3169A>T variant (**Fig. S3F**) and the c.425dupA variant (**Fig. S3H**) also introduce a premature stop codon and trigger NMD as the mRNA level of *JAK1* in the peripheral blood of variant carriers was approximately half that of normal controls. The mRNA level of *JAK1* remains comparable among individuals in pedigree C (**Fig. S3G**).

**Figure 2.**
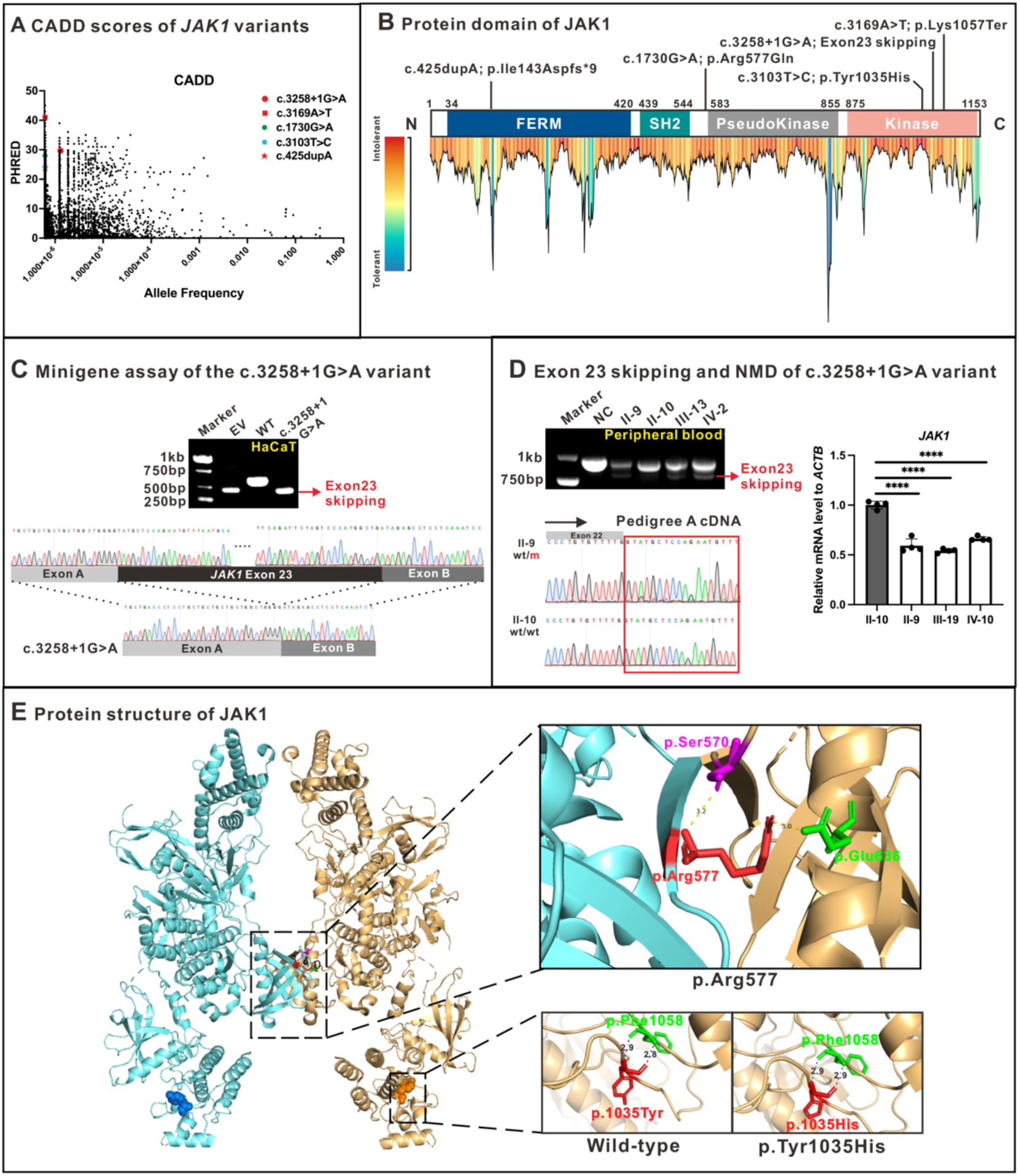
Genetic analyses. (Panel A) Minor allele frequencies for heterozygous *JAK1* variants present in gnomAD (v4.1.0) mapped against corresponding CADD scores (99% confidence interval, HGMD). In the gnomAD database, only one LOF variant exhibits a high frequency: *JAK1*: c.414C>A (p.Tyr138Ter), predicted to introduce a premature stop codon. Notably, this variant is consistently linked with c.412T>C (p.Tyr138His). Collectively, these two variants result in p.Tyr138Gln, resulting in a nonsense-rescued multi-nucleotide variant (MNV). (Panel B) The localization of each variant. The c.425dupA variant maps to the FREM domain, while the c.1730G>A variant is located between the SH2 domain and pseudokinase domain. The c.3103T>C, c.3169A>T, and c.3258+1G>A variants are localized to the kinase domain. Protein tolerance landscape for missense variants in *JAK1* was visualized by MetaDome 1.0.1. The c.1730G>A(p.Arg577Gln) and 3103T>C (p.Tyr1035His) variants in *JAK1* were predicted to be intolerant and highly intolerant, respectively. (Panel C) Minigene assay suggested that the c.3258+1G>A variant leads to exon 23 skipping of *JAK1*. (Panel D) RT-qPCR and Sanger sequencing of blood-derived mRNA in Pedigree A. The c.3258+1G>A variant introduces a premature stop codon and triggers NMD. Accordingly, the mRNA level of *JAK1* in the peripheral blood of variant carriers was approximately half that of normal controls. (Panel E) Three-dimensional structural diagrams of two missense variants. The p.Tyr1035His results in the elongation of the hydrogen bond, increasing its length from 2.8 Å to 2.9 Å. Moreover, Arg577 (corresponding to Arg576 of mouse JAK1) is located at the pseudokinase-pseudokinase dimerization interface of JAK1. The p.Arg577Gln variant may affect the JAK1 PK-PK dimerization process, thereby influencing the downstream phosphorylation^22^. The p.Tyr1035 is one of the major phosphorylation sites of JAK1 and is essential for function.

**Supplementary Figure 3.**
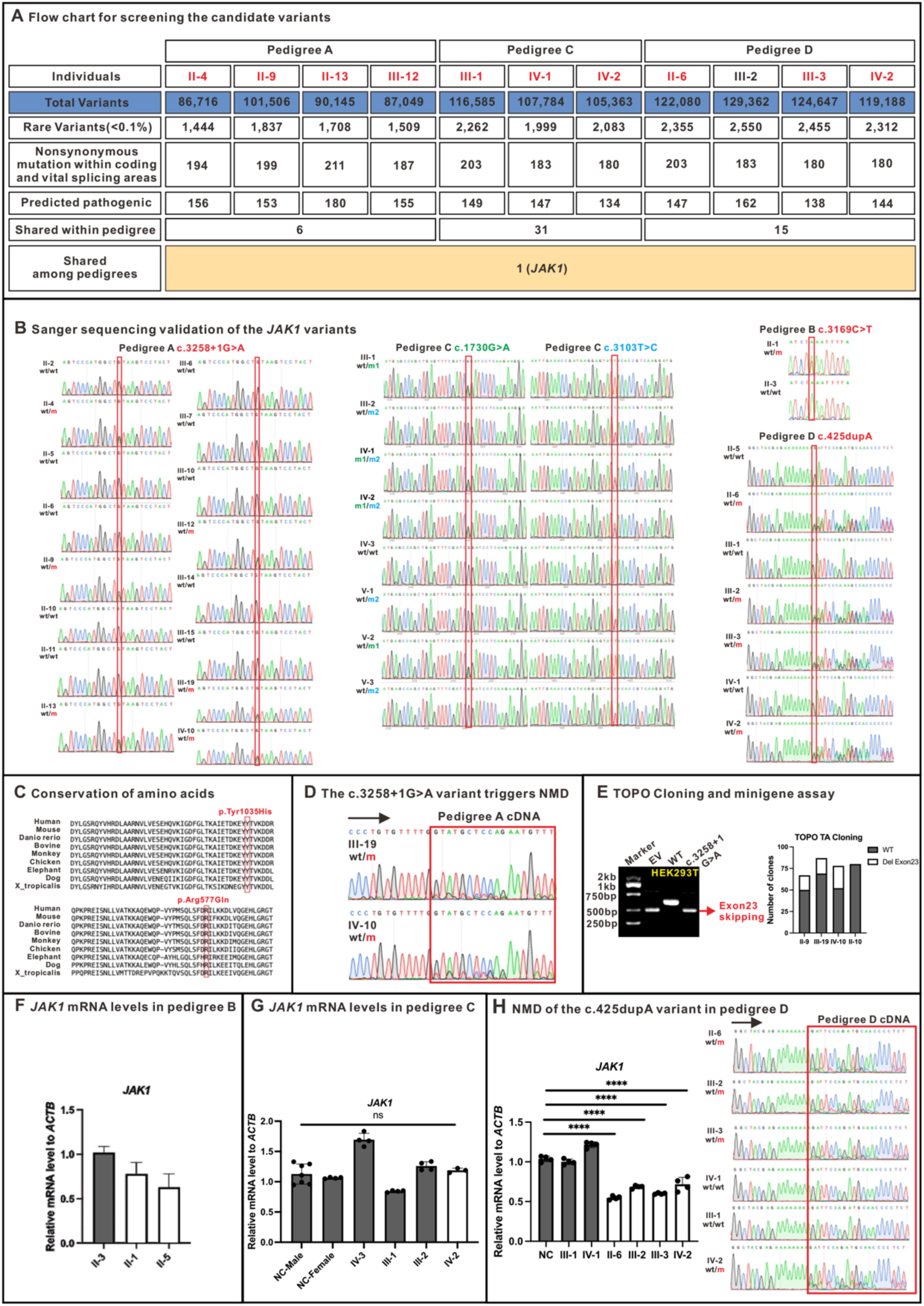
Genetic analyses of *JAK1* variants. (Panel A) Flow chart for screening the candidate variants. A total of 6, 31, and 5 rare variants were found to co-segregate within the pedigrees A, C, and D, respectively. Among the segregating mutated genes in each family, *JAK1* is the only one associated with antiviral immunity. The only candidate gene shared by the three pedigrees was *JAK1*. Given the common incomplete penetrance of EV caused by variants in other genes, all unaffected individuals were excluded from candidate gene screening. (Panel B) Sanger sequencing validation of the *JAK1* variants. (Panel C) Evolutionary conservation analysis of p.Arg577 and p.Tyr1035. These two amino acids are highly conserved across species. (Panel D) The c.3258+1G>A variant leads to exon 23 skipping of *JAK1*, introducing a premature stop codon and triggering NMD. (Panel E) TOPO cloning and minigene assay in HEK293T indicated that the c.3258+1G>A variant leads to exon 23 skipping of *JAK1*, introducing a premature stop codon and triggering NMD. (Panel F, G and H) *JAK1* mRNA expression levels in pedigrees B, C, and D. The black bars represent individuals without pathogenic *JAK1* variants, while the white bars indicate those carrying pathogenic *JAK1* variants. (Panel F) The c.3169A>T variant introduces a premature stop codon and triggers NMD. (Panel G) RT-qPCR suggested that the mRNA level of *JAK1* remains comparable among individuals in pedigree C. (Panel H) The c.425dupA variant introduces a premature stop codon and triggers NMD.

### *JAK1* variants caused a defective JAK-STAT pathway

Whether *JAK1* variants led to defective expression and functional effects was further explored. *JAK1*-KO and *JAK1*-KD cells were generated (**Fig. S4B and Fig. S4C**). When overexpressing *JAK1* variant sequences in HaCaT, HEK293T cells, and *JAK1*-KO HaCaT cells, all the variants lead to a decreased level of the JAK-STAT pathway (**Fig. 3A, Fig. S4A, Fig S5A, B, and C**), indicating *JAK1* LOF. *JAK1* LOF was again verified by dual luciferase assay (**Fig. 3B**). *JAK1*-KO Jurkat decreased the level of the JAK-STAT pathway (**Fig. S5D**) and abolished the transcription of many interferon-stimulating genes (ISGs) (**Fig. S5E**), indicating a defective interferon response and cell activation. Patient samples were further employed to confirm the expression and LOF effects of *JAK1* variants. Compared with the skin of normal controls, the JAK1 expression and the JAK1-STAT pathway were decreased in the SCC tissue of II-9 from pedigree A and IV-2 from pedigree C (**Fig. 3C, 3E, and 3F).** The JAK1-STAT pathway was also decreased in the PBMC of IV-2 from pedigree C at the baseline or upon IFNα and IFNγ stimulation (**Fig. 3D**).

**Figure 3.**
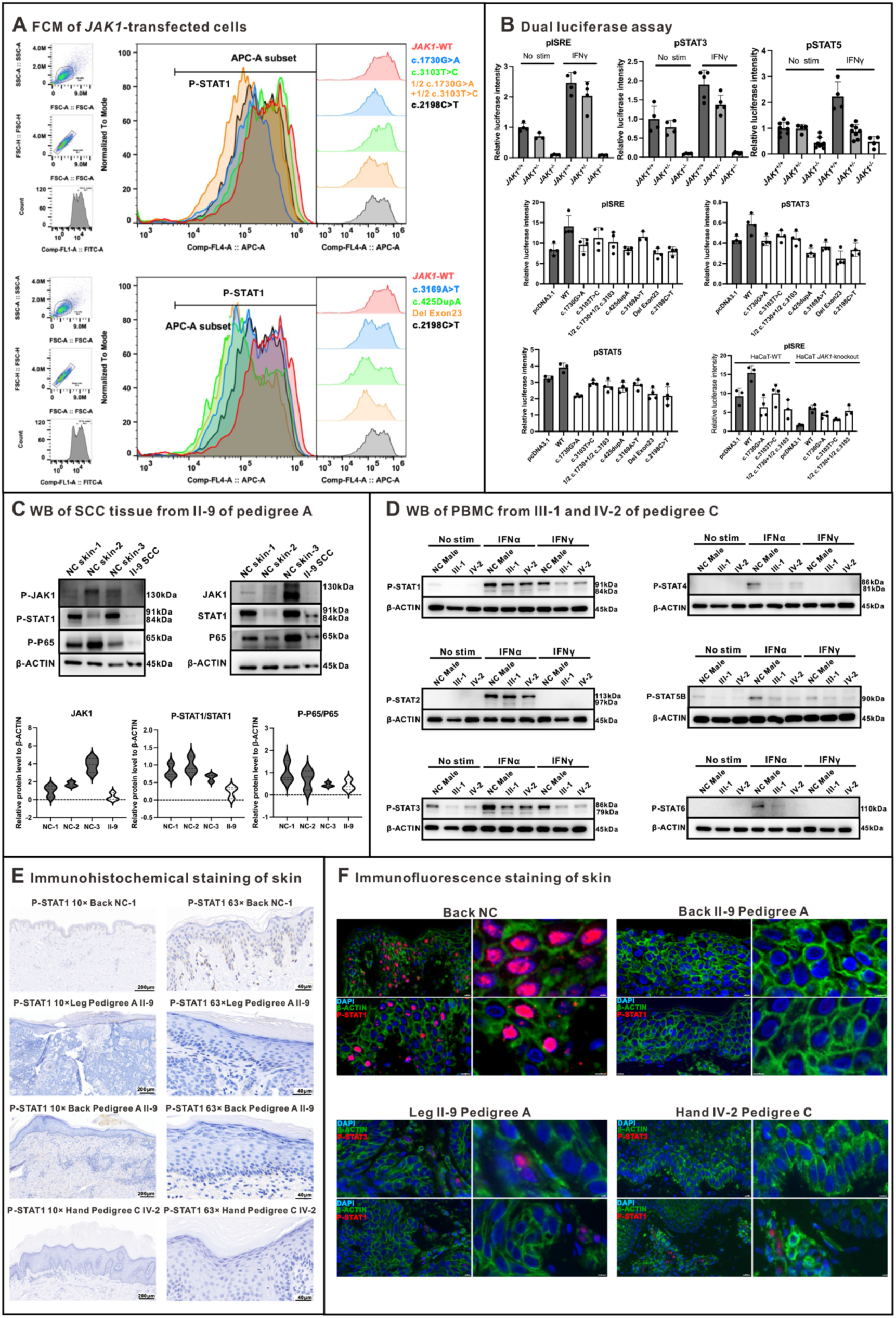
*JAK1* variants caused a defective JAK-STAT pathway. (Panel A) Flow cytometry showed that the JAK-STAT pathway decreased at the baseline or upon stimulation in cells transfected with the *JAK1* variant sequences. c.2198C>T was selected as the *JAK1* LOF positive control. (Panel B) KO and KD of *JAK1* in HaCaT cells reduced, indicating impaired JAK-STAT signaling. HEK293T overexpressing the *JAK1* variants showed decreased pISRE, pSTAT3, and pSTAT5 luciferase activity. Wild-type and *JAK1*-KO HaCaT cells transfected with the c.1730G>A and c.3103T>C variants showed reduced pISRE luciferase activity. (Panel C) The JAK1 expression and the JAK1-STAT pathway were decreased in the SCC skin of II-9 from pedigree A. (Panel D) The JAK1-STAT pathway was decreased in the PBMC of IV-2 from pedigree C at the baseline or upon IFNα and IFNγ stimulation. (Panel E and Panel F) Immunohistochemical staining and immunofluorescence staining indicated that the JAK1-STAT pathway was decreased in the SCC skin of II-9 from pedigree A and IV-2 from pedigree C.

**Supplementary Figure 4.**
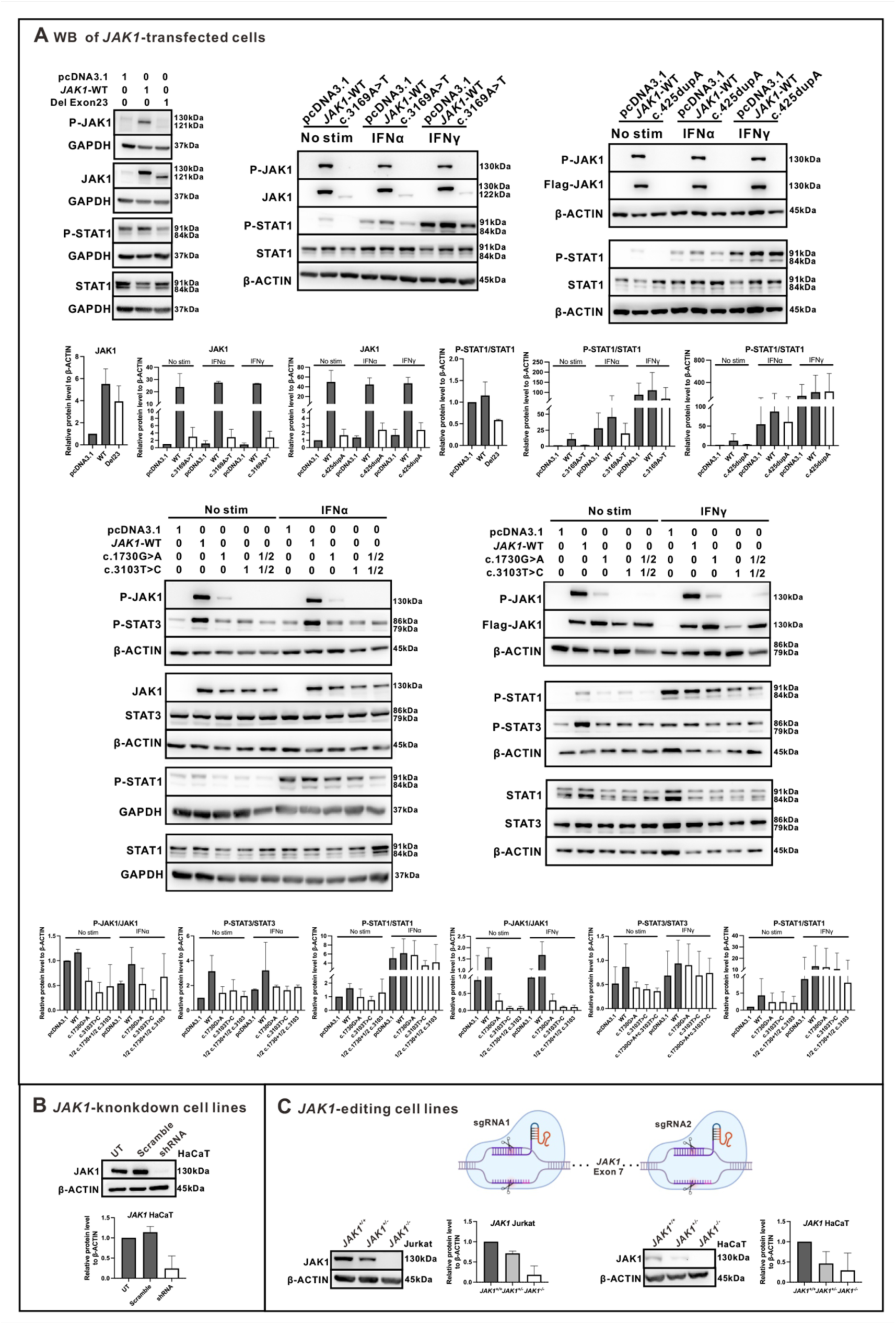
(Panel A) The JAK-STAT pathway decreased at the baseline or upon stimulation in cells transfected with the *JAK1* variant sequences. The c.425dupA variant led to the complete abolition of JAK1 protein expression. The c.3169A>T variant and *JAK1* without exon 23 generated a low level of truncated JAK1 protein. Representative results from three independent replicate experiments. (Panel B) *JAK1* was successfully knocked down in HaCaT. (Panel C) *JAK1* was successfully knocked out in both HaCaT and Jurkat. The sgRNA for CRISPR is targeting Exon 7 of *JAK1*.

### *JAK1* LOF exerts no effect on the known pathogenic mechanism of typical EV

We also investigate whether *JAK1* variants are involved in the known pathogenic mechanism underlying typical EV. Expression of the known EV-causative genes, *CIB1*, *TMC6*, and *TMC8,* was not altered after *JAK1* KD and overexpression of *JAK1* missense variants (**Fig. S6A**). In addition, NF-κB and AP-1 luciferase intensity were not changed when knocking down *JAK1* (**Fig. S6B**). The relative intensity of long control region (LCR) luciferase of the HPV5 and HPV8 was not changed (**Fig. S6B**). The cell proliferation assay demonstrated that neither *JAK1* KD nor *JAK1* KO affects the proliferation rate of cells (**Fig. S6C**). To sum up, *JAK1* LOF exerts no effect on the known pathogenic mechanism underlying typical EV.

**Supplementary Figure 5.**
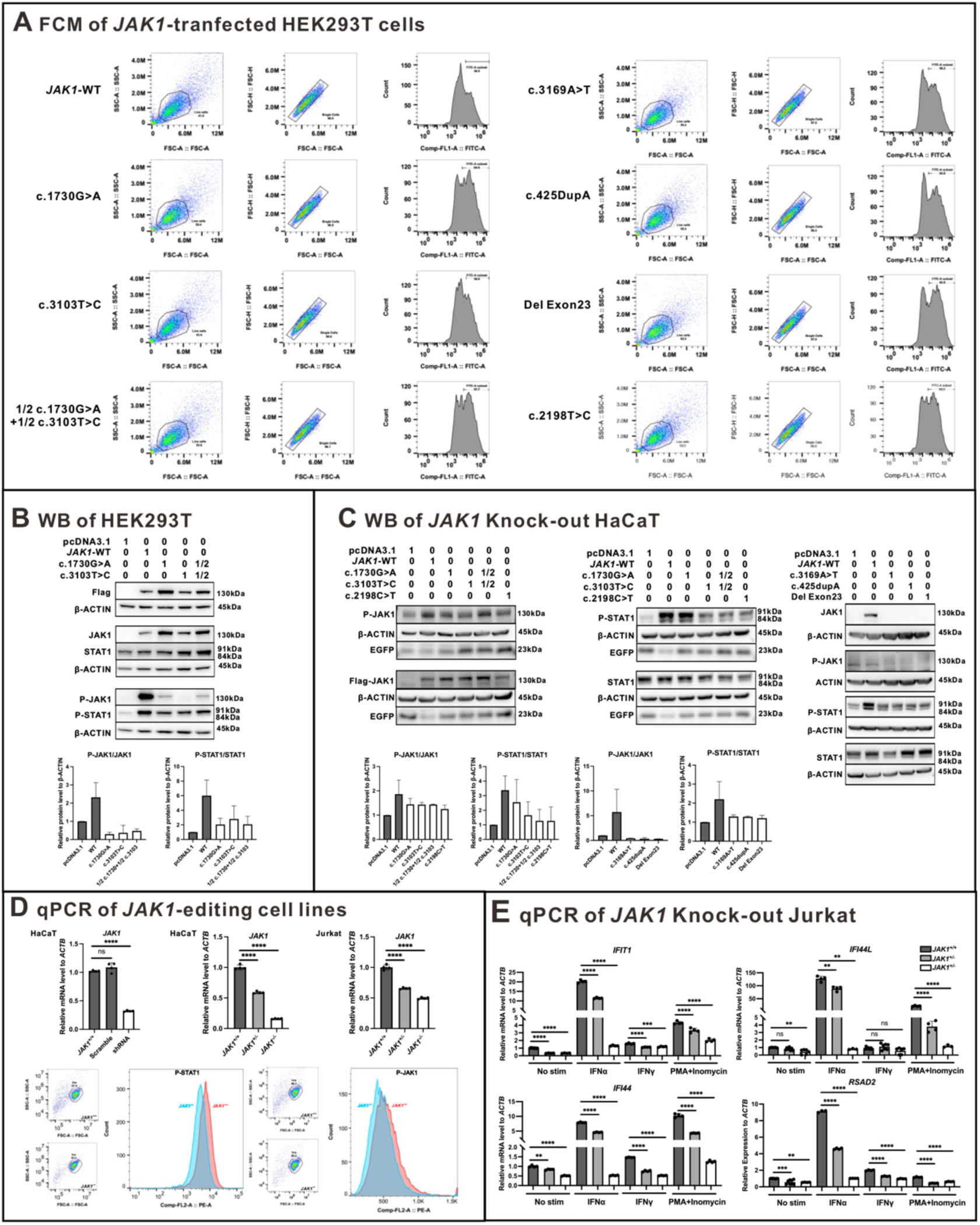
*JAK1* variants caused a defective JAK-STAT pathway. (Panel A) Flow cytometry showed that the JAK-STAT pathway decreased at the baseline or upon stimulation in HEK293T cells transfected with the *JAK1* variant sequences. (Panel B) Western blot showed that the JAK-STAT pathway decreased in HEK293T cells transfected with the *JAK1* variant sequences. (Panel C) Western blot showed that the JAK-STAT pathway decreased in *JAK1*-KO HaCaT cells transfected with the *JAK1* variant sequences. The c.425dupA variant led to the complete abolition of JAK1 protein expression. The c.3169A>T variant and *JAK1* without exon 23 generated a low level of truncated JAK1 protein. (Panel D) The mRNA level of *JAK1* in *JAK1*-KO and *JAK1*-KD cells. The JAK-STAT pathway decreased in *JAK1*-KO Jurkat cells. (Panel E) *JAK1* knockout abolished the transcription of many ISGs upon IFN or PMA/ionomycin stimulation in Jurkat cells.

### A decreased ratio of CD8^+^ naïve T cells was observed in patients

PBMC from different pedigrees were sampled for scRNA-Seq. The T cell subsets were decreased in patients (**Fig. 4A**). The ratio of CD4^+^ T cells/CD8^+^ T cells (**Fig. 4B and Fig. S6D**), especially CD8^+^ naïve T cells (**Fig. 4C and Fig. S6E**), was decreased in patients. The reduction of CD4^+^ and CD8⁺ naïve T cells was also observed in patients with biallelic *JAK1* LOF^17^. Notably, the proportion of CD8⁺ naïve T cells correlates dose-dependently with phenotypic severity in patients and variant carriers (**Fig. 4C**), suggesting its potential role in disease onset and progression.

**Figure 4.**
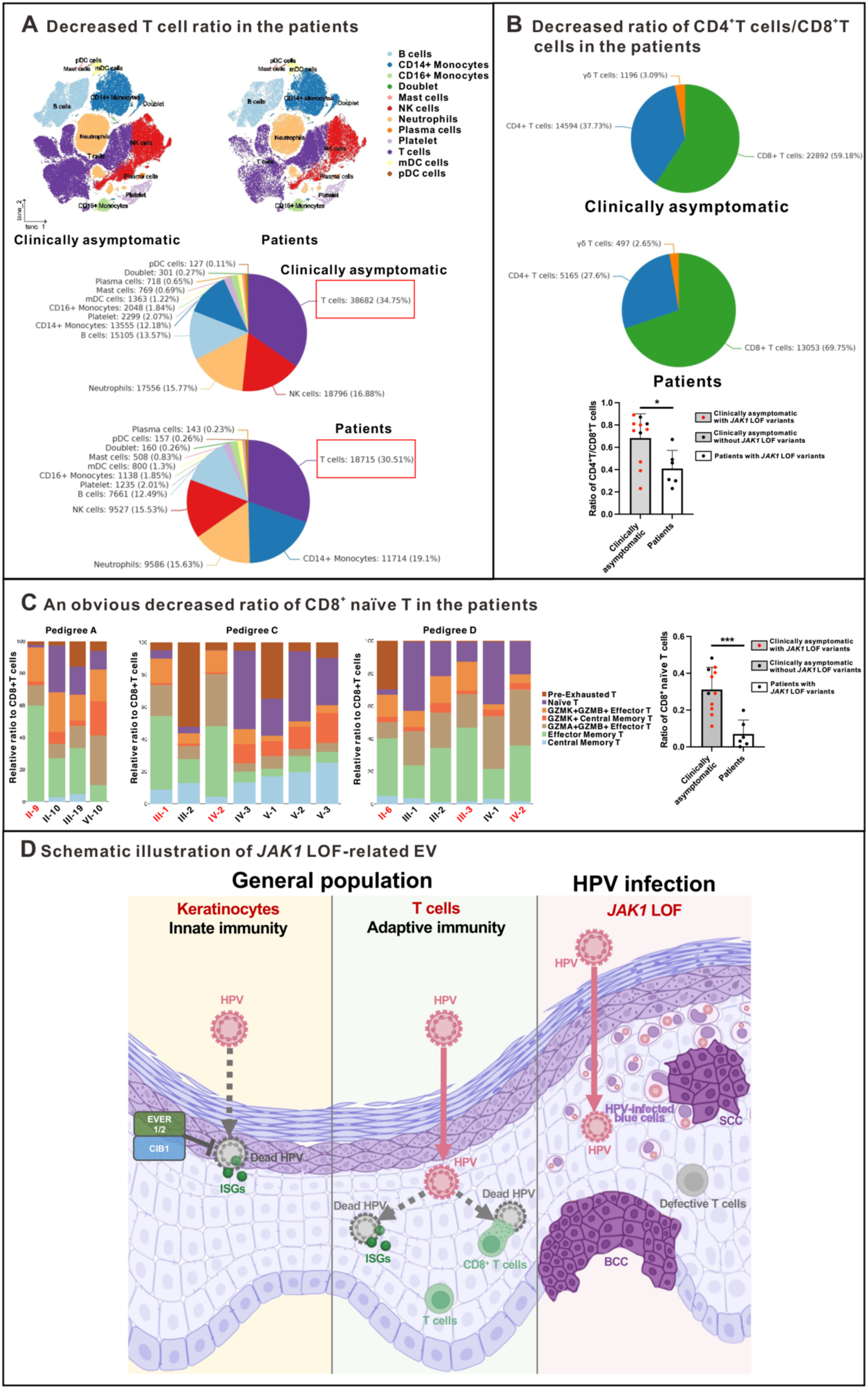
scRNA-Seq revealed immune dysregulation in the peripheral blood and skin lesions of patients with EV. (Panel A) The ratio of T cells was decreased in patients. (Panel B) The ratio of CD4^+^T cells/CD8^+^T cells was decreased in patients. (Panel C) Patients and individuals carrying the *JAK1* variants showed a drastic reduction of naïve CD8^+^T cells. “Patients” means II-9 (Pedigree A), III-1 and IV-2 (Pedigree C), and II-6, III-3, and IV-2 (Pedigree D). “Clinically asymptomatic” indicates II-10, III-19, and VI-10 (Pedigree A), III-2, IV-3, V-1, V-2, and V-3 (Pedigree B), III-1, III-2, and IV-1 (Pedigree D). (Panel D) Schematic illustration of HPV infection associated with *JAK1* LOF. Healthy normal skin can robustly activate T-cell immune responses (green cells) and the expression of ISGs (green dots), which act synergistically to eliminate HPV efficiently (Left: Healthy). In contrast, skin with *JAK1* LOF exhibits defects of T-cell adaptive immunity (gray cells) and impairment of cutaneous innate immunity. HPV persistently proliferates in the skin, characterized by vacuolated cells, and subsequently leads to the development of NMSC (Right: *JAK1* LOF). (Created by BioRender).

**Supplementary Figure 6.**
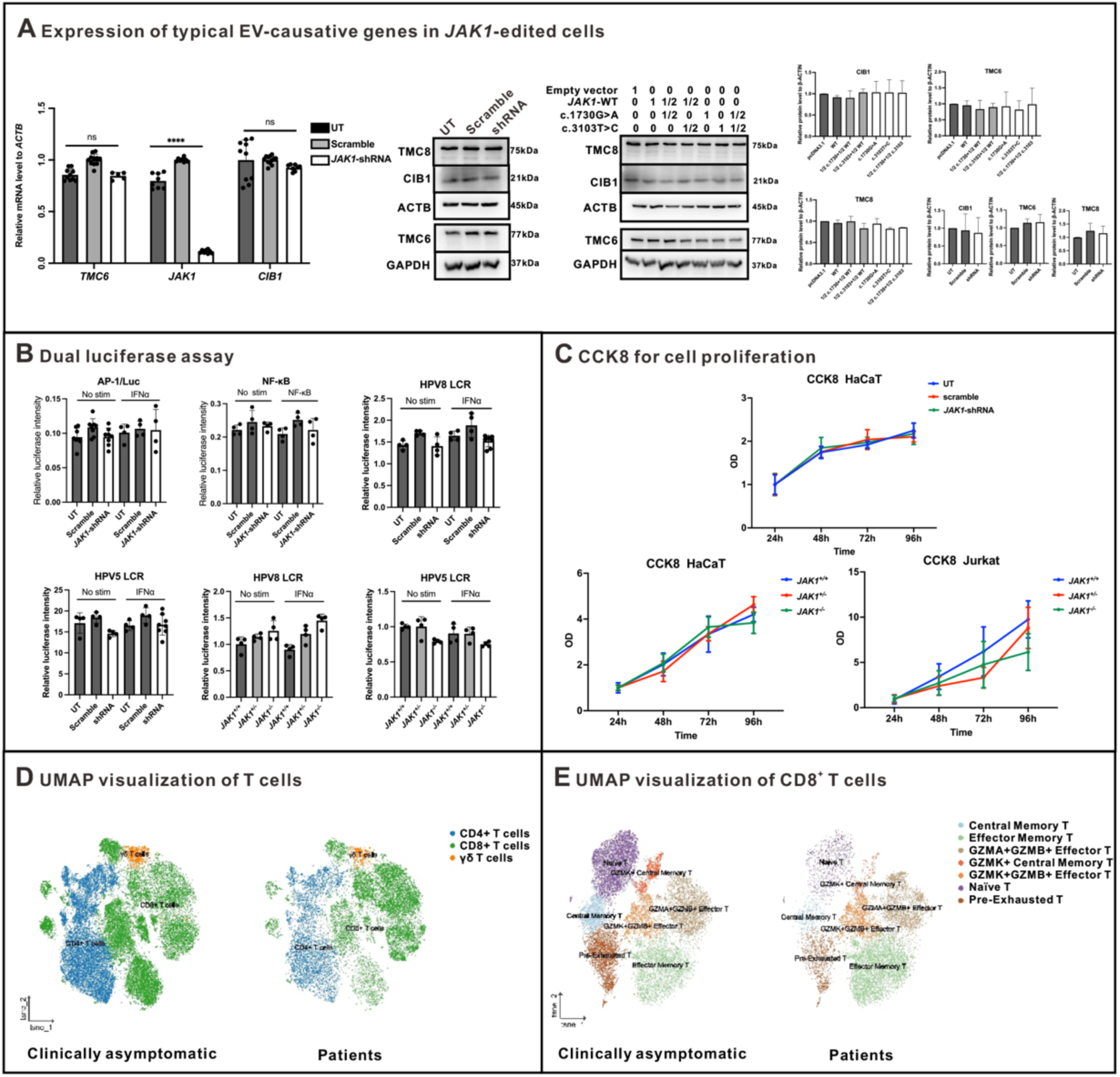
*JAK1* LOF exerts no effect on the known pathogenic mechanism of typical EV. (Panel A) RT-qPCR and Western blot suggest that the expression of the known typical EV-causative genes, *CIB1*, *TMC6*, and *TMC8,* was not altered after *JAK1* knockdown. (Panel B) Dual luciferase assay. NF-κB and AP-1 luciferase intensity were not changed when knocking down *JAK1*. *JAK1* knockdown exerted no effect on the LCR of the HPV. (Panel C) The CCK8 proliferation assay demonstrated that *JAK1* knockout affects neither the proliferation rate of HaCaT cells nor that of Jurkat cells. (Panel D) UMAP visualization of T cells from scRNA-seq data revealed that the ratio of CD4^+^ T cells/CD8^+^ T cells was decreased in the patients. (Panel E) UMAP visualization of CD8^+^ T cells from scRNA-seq data revealed a drastic reduction of naïve CD8^+^ T cells in the patients.

## DISCUSSION

In this study, we identified natural models of human genetic *JAK1* loss with diverse genetic backgrounds and non-consanguineous origins, encompassing both monoallelic and biallelic forms. We reported four independent pedigrees with EV harboring different *JAK1* LOF variants (NM_002227.4, c.3258+1G>A; c.3169A>T; c.425dupA; c.1730G>A, and c.3103T>C). In these pedigrees, different variant forms (missense variants, splicing variants, and indels) collectively lead to *JAK1* LOF. To the best of our knowledge, this is the first time that the relationship between *JAK1* LOF variation and HPV infection has been identified. These *JAK1* variants caused *JAK1* LOF with impaired JAK-STAT pathway and defective anti-viral immunity in both T cells and keratinocytes (**Fig. 4D**). In *Jak1*-KO mice, the number of thymocytes and the development of B cells are severely impaired, and the mouse-derived cells are unresponsive to cytokines using the γc chains^23^. Planar warts restricted to the forehead were also observed in a patient born to consanguineous parents with *JAK1* biallelic LOF variants^17^. However, this phenomenon was not further investigated as the patient exhibited a broader spectrum of immunodeficiency against pathogens. *JAK1* somatic variants are highly prevalent in endometrial cancers^24^, which are also associated with HPV infection^25^. These findings suggest that long-term JAK1 inhibition may be intolerable in humans, with a potential side effect of HPV infection, particularly in skin tissue. In addition, HBV infection was widely observed in our patients, with long-term recurrent infection being particularly prominent. These phenomena may stem from impairment of the interferon pathway induced by *JAK1* LOF. Similarly, double-stranded DNA (dsDNA) virus infections have been reported in patients with EV, such as EBV^26^ and HBV^27^.

Although many EV patients present with severe manifestations, the disorder demonstrates marked phenotypic heterogeneity and incomplete penetrance. Late-onset cases have been reported, with onset as late as 45 years^28^. Therefore, clinically asymptomatic individuals younger than 45 years may have latent HPV infection. III-19 (Pedigree A) and III-2 (Pedigree D), both middle-aged females, had less UV exposure, which might mitigate disease progression. Notably, III-19 from pedigree A showed no overt phenotypes in the original report from two decades ago^29^. In addition, female hormones may act as protective factors, leading to the milder phenotype in A III-12. Vaccination and medication history also influence the phenotype. Although high-risk HPV antibody titers were comparable (all are below the positive threshold) (**Table S8**), EV-associated HPV specific antibodies might differ. The low titers in A III-19 suggest incomplete penetrance was not due to cross-immunity against β-HPV from natural antibodies or vaccination. Decreased CD8^+^ naïve T cell numbers in carriers also suggest latent HPV infection. Individuals with lower T cell proportions, especially CD8+ naïve T cells, are more prone to HPV-related phenotypes. This reduction may be caused by *JAK1* variants and immune senescence due to persistent HPV infection. Furthermore, allele-specific expression in phenotype-relevant cells (e.g., T cells) might lead to skewed *JAK1* allele mRNA levels (as already reported in *JAK1*^30^) and impaired function in cells preferentially expressing the variant allele. However, such cells may undergo cell death, making them unavailable for further analysis. Moreover, genetic modifiers (protective/risk factors, HLA, genetic compensation, transcriptional adaptation) may contribute to reduced penetrance^31^. However, no protective HLA haplotypes were identified (**Table S9**). Somatic reversion and maternal microchimerism may also be protective. Lastly, although no obvious lesions were seen on the face and limbs, pedigree A III-19 had atopic skin reactions, which may partially compensate for the JAK-STAT pathway defect. Pedigree C IV-1 and IV-2 showed much more severe skin lesions, likely due to biallelic LOF variants in *JAK1*. Pedigree C III-2 carried only one variant with milder functional defects and thus remained asymptomatic. The phenotypes are likely shaped by multiple combined factors concertedly.

Typical EV usually arises from the disruption of skin-restricted nonhematopoietic disturbances in keratinocytes^12^. Keratinocytes not only act as physical barriers but also are vital for skin-intrinsic innate immunity^32^. The prominent naïve-memory T cell imbalance in our patient cohort mimics the states of T cell aging^33^, indicating the dysfunction of T cells. Interestingly, HPV16 E7 protein interferes with IFN-γ-mediated JAK1/STAT1 signaling and impairs MHC class I antigen presentation^34^. HPV proteins also suppress STAT1 to support viral genome amplification and episome maintenance^35^, again indicating the important role of the JAK-STAT pathway in HPV infection.

Patients with EV should pay particular attention to protecting their skin from UV exposure. Therapies such as fluorouracil might yield better efficacy in the early stage of the disease course. Hematopoietic stem cell transplantation^36^ and the HPV vaccine^37–39^ have also been reported to be useful in the therapy of HPV-associated skin cancer. Considering the patient exhibits intact antibody-producing capacity, the HPV vaccine might be effective in *JAK1*-related patients with EV.

The nature of this host-virus interplay remains largely unknown. Therefore, EV is an extreme form and a valuable model for investigating cutaneous immunity to HPV and NMSC development. Since some low-frequency *JAK1* variants with high CADD scores were identified in the gnomAD database, certain patients with HPV infection or mild phenotypes might have been overlooked, which aligns with reports that some rare diseases can be masked by molecular phenotypes that are not observed^31,40^. Our work highlights *JAK1*-dependent signaling as a key pathway for defense against HPV. The identification of patients with rare *JAK1* LOF variants provides a natural model for understanding the long-term human response to JAK1 inhibition, and also emphasizes the attribution of *JAK1* deficiency to HPV infection. Our findings also offer a core basis for re-evaluating the safety thresholds and adverse effect risks of HPV infection of JAK1-targeted therapies.

## ETHICS STATEMENTS

Clinical information and blood samples were collected from the family trio, and all participants (or legal guardians if the patient was a minor) signed the informed consent form. This study was approved by the Peking Union Medical College Institutional Review Board in accordance with the Declaration of Helsinki (Approval No: zs2024016).

## DATA AVAILABILITY STATEMENT

All datasets are included in the manuscript or in the supplementary material.

## AUTHOR CONTRIBUTIONS

Xue Zhang and Dong-Lai Ma designed the whole study. Clinical samples and professional medical guidance were provided by Roberto Colombo, Alessandro Pontoglio, Hong Liu, Zhi-Miao Lin, Fu-Ren Zhang, Jia-Wei Liu, and Dong-Lai Ma. Shi-Qi Fan, Rong-Rong Wang, Kai-Chen Tang, Li-Li Zhang, Kai Li, Shi-Rui Han, Han Zhang, Xiao Bai, Xue Yu, Xiaerbati Habulieti, Ke-Qiang Liu, Yang Sun, Li-Wei Sun, and Jia-Cheng Li conducted all the experiments. Miao Sun and Xue Zhang supervised the progress. Shi-Qi Fan, Rong-Rong Wang, Roberto Colombo, and Kai-Chen Tang prepared the first draft of the manuscript. Shiqi Fan, Kai-Chen Tang, Miao Sun, and Xue Zhang independently checked and revised the syntax mistakes and logical errors. All the authors listed contributed to the manuscript and approved the final version to be submitted.

## ACKNOWLEDGMENTS

We’d like to thank all the patients and clinicians for their generous support and collaboration.

