## Supplementary Appendix 1 for "Genetic loss of *JAK1* and cutaneous HPV infection"

**Whole-exome/genome sequencing based on family trio (Trio-WES/WGS)**

Genomic DNA was extracted from the peripheral blood of the family trio using a QIAamp DNA Blood Mini Kit (QIAGEN, Cat. No. 51106), excluding pedigree D II-5. For pedigree D II-5, DNA was extracted from five sterile throat swabs (TIANGEN Biotech DP322-02). The viral DNA was extracted from formalin-fixed, paraffin-embedded (FFPE) SCC tissues obtained from the probands in pedigree C and D (TIANGEN Biotech, Cat. No. DP331-02). The viral DNA from the proband in pedigree A was extracted from the SCC tissue. For individuals III-1 and IV-2 in pedigree C, and II-6 in pedigree D, viral DNA was isolated from lesional skin and adjacent normal skin using separate sets of five sterile swabs each (TIANGEN Biotech, Cat. No. DP322-02), with normal skin serving as the control. DNA libraries were sequenced using an Illumina Novoseq X plus platform with a 100× read depth (WES) or 30× read depth (WGS).

**Variant screening**

Candidate pathogenic variants were further screened. Firstly, the variants in the known EV-causative genes (*ADA*, *ADA2*/*CECR1*, *ARPC1B*, *ATM*, *ATP2C1*, *CARD11*, *CARMIL2*/*RLTPR*, *CD3D*, *CD28*, *CD40LG*, *CF*, *CIB1*, *CORO1A*, *CXCR4*, *DCLRE1C*, *DOCK8*, *FANCA*, *FANCC*, *FANCD2*, *FANCF*, *FANCI*, *GATA2*, *HRAS*, *ICOS*, *IKBKG*/*NEMO*, *IL2RG*, *IL7*, *IL7R*, *ITK*, *JAK3*, *LIG4*, *MAGT1*, *NFKB1*, *NLRP1*, *PIK3CD*, *PTEN*, *SASH3*, *SPINK5*, *STK4/MST1*, *TMC6/EVER1*, *TMC8/EVER2*, *WAS*, *CD4*, *NF1*, *NFKBIA*, *RFXANK*, *TPP2*, *ZAP70*, *ANKRD26*, *CARD9*, *CASP10*, *CD27*, *CHUK*, *DNMT3B*, *FAT4*, *FCGR3A*/*CD16*, *HYOU1*, *ICOSLG*, *IFNGR2*, *IKZF3*/*AIOLOS*, *IRF8*, *ITGB2*/*CD18*, *IVNS1ABP*, *LCK*, *LRBA*, *MALT1*, *MR1*, *MYD88*, *MYH7*, *PGM3*, *PIK3R1*, *POLD1*, *PTPRC/CD45*, *RAC2*, *RAG1*, *RHOH*, *RASGRP1*, *RBCK1*/ *HOIL1*, *SMARCAL1*, *STAT1*, *STAT3*, *TNFSF12*/*TWEAK*, *TRAC*, *FLT3R*, *ITPR3*, *NFATC1*, *FLT3L, ITGAL*) were checked. WGS were performed on the probands of Pedigree A, C, and D for excluding noncoding and structural variants in these genes. After excluding common variants (frequency no less than 0.1% in the Genome Aggregation Database (gnomAD v2.1.1), Exome Aggregation Consortium database（ExAC）, 1000 gnomes project database, and NHLBI-ESP project database database), no explainable pathogenic variants were found. Subsequently, novel candidate pathogenic variants were screened using WES data. Variants with a sequencing depth of at least 20 were retained for quality control. Assuming that pedigree A, C, and D displayed AD and XD inheritance, common variants with a frequency of more than 0.1% in the public databases were removed. We then retained nonsynonymous variants located within exons or those predicted by SpliceAI to impact splicing. Next, variants predicted to be pathogenic by at least one of SIFT, Polyphen, MutationTaster, and CADD v1.7 were retained. All the candidate variants were validated by Sanger sequencing. The variant frequency was further confirmed to be less than 0.001 in gnomAD v4.1.0. Primers for Sanger sequencing were listed in **Table S1.**

**Bioinformatic analyses and molecular modeling**

MutationTaster, InterVar, FATHMM_MKL, CADD v1.7, SIFT, Polyphen2, SpliceAI, and varSEAK Online were used to predict the pathogenicity of the *JAK1* variants. The JAK1 protein sequences of each species were downloaded from UniProt and aligned via CLUSTALW. The cryo-EM model of JAK1 dimer is downloaded in the Protein Data Bank (PDB) under accession code 7T6F. The structures were visualized by PyMOL best rotamers were selected for the coordinates. Tolerance landscape for missense variants in *JAK1* was predicted and visualized by MetaDome 1.0.1 (https://stuart.radboudumc.nl/metadome/dashboard).

**Cell culture, plasmids, and transfection**

HEK293T and HaCaT were maintained in Dulbecco's Modified Eagle Medium supplemented with L-glutamine (Gibco™, Cat. No. 25030081), Jurkat was maintained in RPMI 1640 (Gibco™, Cat. No. 11875093) supplemented with L-glutamine (Gibco™, Cat. No. 25030081). These cell culture media were supplemented with 10% fetal bovine serum (Gibco™, 10091148) and 1% penicillin‒streptomycin (Gibco™, Cat. No. 15140163). The small-interfering RNA for constructing the *JAK1* knock-down (KD) cell line was: 5’-GCATGGAACCAACGACAATGATTCAAGAGATCATTGTCGTTGGTTCCATGCTTTTTT-3’, and the scramble small-interfering RNA was: 5’-GCACCCAGTCCGCCCTGAGCAAA-3’. The single guide RNA (targeting exon 7) used for constructing the *JAK1* knock-out Jurkat cell line was gRNA 1: 5’-TGACCATCATAAGGAGATGC-NGG-3’ and sgRNA 2: 5’-AGGAACTTAACCTAGATCTC-NGG3’. The guide RNA (targeting exon 7) used for constructing the *JAK1* knock-out (KO) HaCaT cell line was sgRNA 1: 5’-TAACTCCATCTAACGTCCTC-NGG -3’ and sgRNA 2: 5’-ACTAGTTAAGTAATTGGCAG-NGG -3’. *JAK1* wild-type and variant sequences were linked into Flag-pcDNA3.1-HA-PKG-EGFP vectors. The transfection was performed using Lipofectamine^TM^ 3000 (Invitrogen, Cat. No. L3000015) according to the manufacturer’s instructions. For western blot, HaCaT cells were stimulated with IFN-α (Sino Biological 12341-H08Y, 1000 IU/mL, 15 min), IFN-γ (Biolegend 570206, 1 ng/mL, 15 min). For reverse transcription quantitative PCR (qRT-PCR), Jurkat cells were stimulated with PMA (Sigma P1585, 100μg/mL, 20 h) together with inomycin (Sigma I3909, 100 μM, 20 h), or CD3 (eBioscience™ 16-0037-81, 0.5 μg/mL, 15 h) together with CD28 (eBioscience™, 14-0289-82 0.5 μg/mL, 15 h). For dual luciferase assay, HaCaT cells were stimulated with IFN-γ (Biolegend 570206, 1 ng/mL, 24 h).

**Protein extraction and western blot**

Cells were washed with 1×PBS (Gibco™, Cat. No. 20012027) and lysed in radioimmunoprecipitation assay (RIPA) buffer (Beyotime, Cat. No. P0013C) containing protease inhibitors (Roche, Cat. No. 5892970001) and phosphatase inhibitors (Roche, Cat. No. 4906837001), and then incubated on ice for 30 min. Then, the cell debris was removed after centrifugation at 4°C, 14,000×g for 20 min. Protein from the whole blood was extracted according to the instructions of the manufacturer (Solarbio, EX1200). The protein lysate was diluted by adding 4× Protein SDS-PAGE Loading Buffer (TAKARA, Cat. No. 9173) and RIPA. Afterward, the mixture was boiled at 98°C for 10 min and then incubated on ice for 2 min. Electrophoresis was performed with 10% TGX Stain-Free polyacrylamide gels (Bio-Rad, Cat. No. 1610182). Then, the protein was transferred into a 0.45 μm Immobilon®-P PVDF Membrane (Millipore, Cat. No. IPVH00010). Subsequently, the membrane was blocked with 1×Tris-buffered saline–Tween 20 (TBS-T) buffer (Solarbio, Cat. No. T1080) containing 5% skim milk (Solarbio, Cat. No. D8340) or 5% Bovine serum albumin (Sigma, Cat. No. V900933) at room temperature for 60 min and incubated with primary antibodies at 4°C overnight for 12 hours to 16 hours. The membrane was washed with 1× TBS-T and then incubated with horseradish peroxidase-conjugated sheep anti-mouse IgG (ZSGB-Bio, Cat. No. ZB-2305) or sheep anti-rabbit IgG (ZSGB-Bio, Cat. No. ZB-2301) at room temperature for 60 min. After another wash with 1× TBS-T, the membranes were exposed using Tanon™ High-sig ECL Western Blotting Substrate (Tanon, Cat. No. 180-5001). WB stripping buffer (TAKARA, T7135A) was used for antibody stripping. The primary antibodies used are listed in **Table S2**.

**RNA extraction and reverse transcription quantitative PCR (RT-qPCR)**

According to the manufacturer's instructions, RNA was extracted using the TRIzol^TM^ Reagent (Invitrogen, Cat. No. 15596018) or TRIzol^TM^ LS Reagent (Invitrogen, Cat. No. 15596018). cDNA was synthesized using a PrimeScript™ RT Master Mix (Perfect Real Time) (TAKARA, Cat. No. RR036A). RT-qPCR was performed using Hieff® qPCR SYBR Green® Master Mix (Low Rox Plus) (Yeasen, Cat. No. 11202ES08) on a Thermo Applied Biosystems™ QuantStudio™ 3 Real-Time PCR System. Human *ACTB* was used as an endogenous control, and the relative gene expression was normalized to that of Human *ACTB* and calculated using the 2^−ΔΔCt^ method. Primers used for RT-qPCR are listed in **Table S1**. The data were presented as the mean ± standard deviation (SD).

**Flow cytometry**

Following washing and lysis, the cells were resuspended in a single-cell suspension and fixed for 30 min (eBioscience™ Intracellular Fixation & Permeabilization Buffer Set, Invitrogen 88-8824-00). For intracellular staining, antibodies were diluted in permeabilization buffer (eBioscience™ Intracellular Fixation & Permeabilization Buffer Set, Invitrogen 88-8824-00) and incubated with the cells in the dark for 60 min. After washing, the samples were analyzed using a BD Accuri™ C6 Plus flow cytometer, and the data were analyzed using FlowJo version 10.8.1. The antibodies used for flow cytometry are listed in **Table S2.**

**Dual luciferase assay**

The sequences of HPV5 long control region (LCR) and HPV8 LCR were constructed according to the HPV genome database (PaVE: the papillomavirus episteme, https://pave.niaid.nih.gov/search/search_database) and incorporated into the multiple cloning site in the pGL4.20 vector. The AP-1/Luc vector was constructed according to a previous reoprt^19^. The NF-κB responsive element pGL4.32 [luc2P/NF-κB-RE/Hygro] was used to assess the NF-κB level. The pISRE-TA-Luc, pSTAT3-TA-Luc, and pGL4.52[luc2P STAT5 RE Hygro] plasmids were used to assess the JAK-STAT pathway. pRL-TK (Renilla luciferase) was chosen as an endogenous control. Luciferase activity was measured on a Synergy H1 reader (BioTek) using the Dual-Luciferase Reporter Assay System (Promega, E1960), and the data were presented as the Firefly luciferase intensity normalized to the Renilla luciferase intensity.

**HBV DNA, HBV Antibody, and interleukins quantification**

The blood samples from patients were used to detect HBV DNA and HBV antibodies. Specifically, HBV DNA was quantified via quantitative polymerase chain reaction (qPCR), and HBV antibodies were measured using Chemiluminescent Immunoassay (CLIA). Cytokine profiles in patient serum samples were determined via CLIA.

**Immunofluorescence staining**

The tissue sections of SCC (II-9 from pedigree A and II-2 from pedigree C) were washed with 1×PBS and fixed with 4% Paraformaldehyde (Sigma, 158127-5G) diluted in 1×PBS. Subsequently, cells were permeabilized by 0.1% Triton-100 (Solarbio, P1080) diluted in 1×PBS and blocked by the mixture of 1% BSA (Sigma, V900933) and 5% normal goat serum (Jackson ImmunoResearch, AB_2336990) diluted in 1×PBS at room temperature for 60 min. Then, tissues were stained using primary antibodies overnight and incubated with Alexa Fluor™ 488nm or Alexa Fluor™ 594nm Antibody. Finally, the cell nucleus was stained with DAPI (10 μg/mL) (Sigma-Aldrich, D9542) for 10 min. Images were captured on Pannoramic MIDI (3DHISTECH) and visualized by CaseViewer2.4 (3DHISTECH). The primary antibodies used for Immunofluorescence staining are listed in **Table S2**.

**Hematoxylin and eosin (H&E) staining and immunohistochemical staining**

The tissue sections of SCC (II-9 from pedigree A and II-2 from pedigree C) were dehydrated gradually. Skin paraﬃn sections were cut into 5 µm-thick sections, dewaxed in xylene, and rehydrated through a descending series of ethanol concentrations. H&E staining was performed according to the manufacturer’s instructions (Solarbio, G1120). Briefly, skin sections were rehydrated, stained with hematoxylin for 5 min, rinsed with tap water, differentiated in 1% acid ethanol for 30 s, rinsed with tap water for 30 min, stained with eosin for 1.5 min, and finally washed with tap water. For immunohistochemical staining, skin sections were dewaxed and rehydrated, followed by antigen retrieval with 1× sodium citrate antigen retrieval solution (Solarbio, C1032) and blocking at room temperature for 1 hour. Primary antibodies diluted in 3% BSA were incubated with sections overnight at 4°C. The next day, sections were sequentially washed with PBST and 1× PBS, then incubated with goat anti-rabbit IgG-Biotin secondary antibody (3% BSA-diluted) at room temperature for 1 hour. After washing, streptavidin-biotin-peroxidase complex was added and incubated for 1 hour at room temperature. Freshly prepared 3,3'-diaminobenzidine was used for dark color development at room temperature. Sections were subsequently counterstained with Harris hematoxylin for 2 min, differentiated with hydrochloric acid-ethanol for 10 seconds, rinsed with running tap water for 10 min, then dehydrated, cleared with ethanol and xylene sequentially, and mounted (Sigma, P-1585). Images were captured on Pannoramic MIDI (3DHISTECH) and visualized by CaseViewer2.4 (3DHISTECH). The primary antibodies used for immunohistochemical staining are listed in **Table S2**.

**Genotyping of HPV**

For library construction, 1 μg genomic DNA was randomly fragmented into ~350 bp segments via Covaris ultrasonic disruptor. Library fragment integrity was assessed by AATI analysis. Clean data were aligned to a host database using Bowtie2 to filter host-derived reads, then assembled with MEGAHIT. Scaffolds were split at N junctions to generate N-free scaftigs. ORFs were predicted by MetaGeneMark from scaftigs (≥500 bp), with sequences <100 nt filtered out. Redundancy was removed using CD-HIT to obtain a non-redundant initial gene catalog. Clean data from each sample were aligned to this catalog via Bowtie2; genes with ≤2 reads per sample were filtered out to confirm the final Unigene catalog. Gene abundance was calculated based on aligned read counts and gene length, followed by basic statistics, core-pan gene analysis, and sample correlation analysis. Unigenes were finally aligned to the PaVE database for HPV typing.

**Cell proliferation assay**

Cell Counting Kit-8 (CCK-8) was used to measure the proliferation rate of cells according to the manufacturer's instructions (DONJINDO CK04, Japan). Briefly, cells were seeded in 24-well plates, with absorbance measured at 24 h, 48 h, 72 h, and 96 h post-seeding.

**Single-cell RNA sequencing (scRNA-Seq)**

Single-cell suspensions were processed through the 10x Genomics Chromium Controller (10x Genomics). The libraries were constructed following the protocol outlined in the Chromium Single Cell 3’ Reagent Kits v3.1 (10x Genomics, 1000268) User Guide. Cell count and viability were estimated using a Cell Analyzer (Countstar® Rigel S2). An estimated 10,000 labeled live cells per sample were loaded into the chip to generate single-cell droplets. Following reverse transcription, the cDNA was generated and amplified. The libraries were pooled for sequencing using an Illumina Novaseq X-25B sequencer with a pair-end 150 bp reading strategy. All samples were assessed for doublet content using DoubletFinder version 2.0.3, and cells called as doublets were removed before further analysis. Meanwhile, cells with fewer than 200 genes, cells with mitochondrial gene context exceeding 25%, and genes expressed in less than 3 cells were filtered out. A linear dimensional reduction was performed, presenting as Uniform Manifold Approximation and Projection (UMAP) or t-distributed Stochastic Neighbor Embedding (t-SNE). Marker genes for scRNA-Seq are listed in **Table S3**.

**Statistical analyses**

The statistical analyses were performed using GraphPad Prism 9.5.1. When the data satisfied both an approximate normal distribution and a homogeneity of variance, a standard t-test was employed for analysis. A Welch’s t-test was performed if the data only satisfied approximate normality. The Mann–Whitney U test was applied for data deviating from a normal distribution. Statistical significance was defined as *p*<0.05. ns, ∗, ∗∗, ∗∗∗, and ∗∗∗∗ indicate not statistically significant, *p*<0.05, *p*<0.01, *p*<0.001, and *p*<0.0001, respectively.
